# Estimating effects of intervention measures on COVID-19 outbreak in Wuhan taking account of improving diagnostic capabilities using a modelling approach

**DOI:** 10.1101/2020.03.31.20049387

**Authors:** Jingbo Liang, Hsiang-Yu Yuan, Lindsey Wu, Dirk U. Pfeiffer

## Abstract

**Background:** Although by late February 2020 the COVID-19 epidemic was effectively controlled in Wuhan, China, the virus has since spread around the world and been declared a pandemic on March 11. Estimating the effects of interventions, such as transportation restrictions and quarantine measures, on the early COVID-19 transmission dynamics in Wuhan is critical for guiding future virus containment strategies. Since the exact number of COVID-19 infected cases is unknown, the number of documented cases was used by many disease transmission models to infer epidemiological parameters. However, this means that it would not be possible to adequately estimate epidemiological parameters and the effects of intervention measures, because the percentage of all infected cases that were documented changed during the first 2 months of the epidemic as a consequence of a gradually increasing diagnostic capability.

**Methods:** To overcome the limitations, we constructed a stochastic susceptible-exposed-infected-quarantined-recovered (SEIQR) model, accounting for intervention measures and temporal changes in the proportion of new documented infections out of total new infections, to characterize the transmission dynamics of COVID-19 in Wuhan across different stages of the outbreak. Pre-symptomatic transmission was taken into account in our model, and all epidemiological parameters were estimated using Particle Markov-chain Monte Carlo (PMCMC) method.

**Results:** Our model captured the local Wuhan epidemic pattern as a two-peak transmission dynamics, with one peak on February 4 and the other on February 12, 2020. The impact of intervention measures determined the timing of the first peak, leading to an 86% drop in the R_e_ from 3.23 (95% CI, 2.22 to 4.20) to 0.45 (95% CI, 0.20 to 0.69). An improved diagnostic capability led to the second peak and a higher proportion of documented infections. Our estimated proportion of new documented infections out of the total new infections increased from 11% (95% CI 1% - 43%) to 28% (95% CI 4% - 62%) after January 26 when more detection kits were released. After the introduction of a new diagnostic criterion (case definition) on February 12, a higher proportion of daily infected cases were documented (49% (95% CI 7% - 79%)).

## Introduction

Coronavirus disease 2019 (COVID-19), an acute respiratory infection originally identified in the city of Wuhan in Hubei Province, China, has spread worldwide in 2020^1-2^. Estimating the effects of intervention measures is still one of the major scientific goals in order to identify proper prevention measures in the world^3^. The precise estimation of transmissibility R_e_ is critical for the identification of appropriate intervention measures to contain the outbreak^1,4,5,6,7,8^. Although many recent studies have evaluated how intervention measures implemented in Wuhan reduced disease spreading to regions outside Wuhan^6,9,10,11,12^, the investigation of the contribution of interventions within Wuhan, the epidemic source region itself, has not been done much^13 14^, possibly because that an irregular pattern of transmission dynamics during early February hinders the model fitting processes, making the precise estimation of the parameters difficult.

To control the virus spreading during the early outbreak stage, the Chinese government implemented strict travel restrictions on January 23, 2020 in Wuhan ^15^. The first epidemic peak occurred twelve days after the restrictions were implemented. Soon afterwards, the number of new daily documented cases started to fluctuate for about two weeks around this peak value, with another extremely high number of cases peaked in the middle, and then finally reduced (Figure S1). The transmission dynamics with such an irregular and unusual pattern can affect the estimation of the effects of intervention measures. The high number of documented cases after the introduction of interventions was generally hypothesized to be mainly caused by improved diagnostic capability^16^, leading to more detected cases rather than caused by the intrinsic growth of the epidemic. However, most studies have not considered the changes in diagnostic capability over time, which can affect the number of documented infections and, ultimately, the estimation of R_e_.

Accounting for temporal changes in COVID-19 diagnostic capability is critical for characterizing transmissibility and understanding the pattern of the local Wuhan epidemic. Recent studies have shown that the total potential case number has been significantly underestimated, with more than 80% of all infections undocumented during the initial period following the identification of SARS-CoV-2 as the causative agent^17^. While the number of total new infections is driven by the epidemic growth, after the introduction of new commercial kits^18^ and introduction of more sensitive diagnostic criteria^16^ (Figure 1), diagnostic capacity in Wuhan increased, resulting in a higher proportion of total new infections been documented. Therefore, it is important to consider the improvements in diagnostic capacity over time when using the documented data to reconstruct transmission models for COVID-19 in Wuhan.

**Figure 1.**
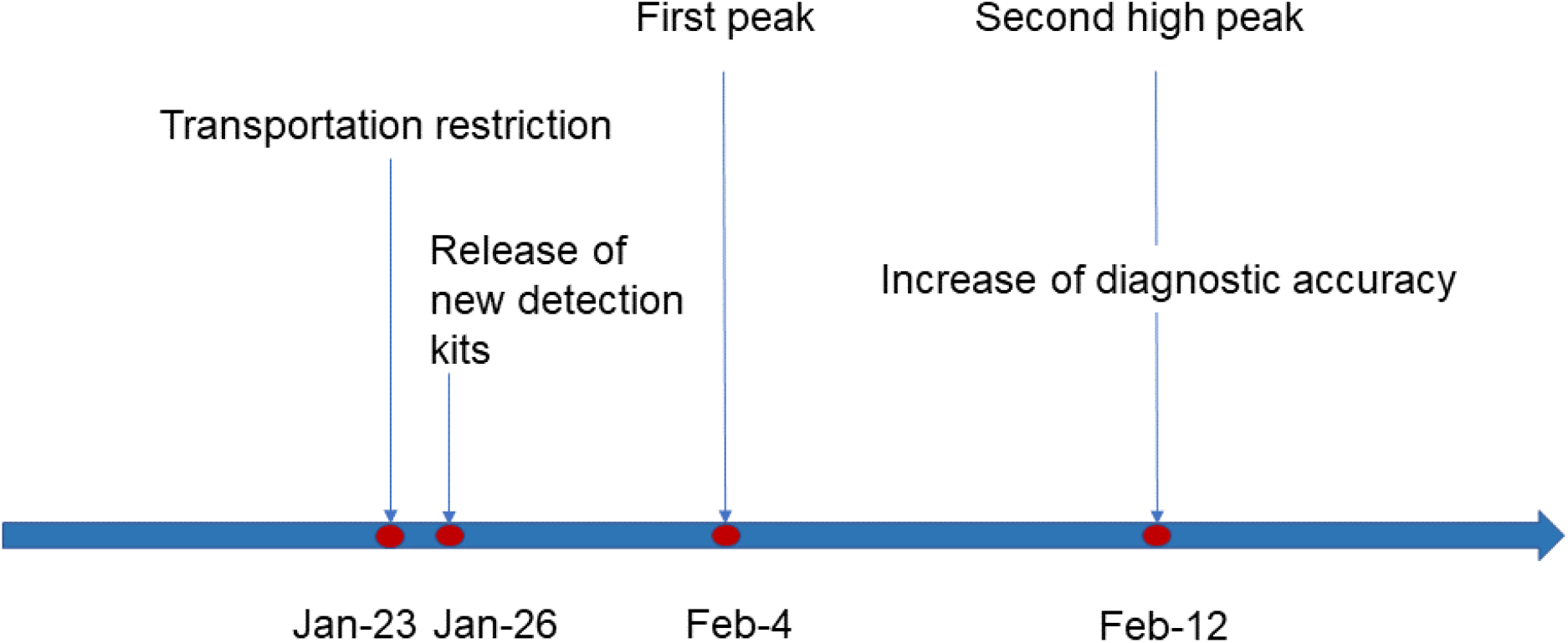
The timeline of improved diagnostic capability and intervention measures implemented in Wuhan, China. New commercial kits were approved by the State Food and Drug Administration (SFDA) on January 26^18^; Updated diagnostic criteria, COVID-19 case confirmation should rely on both clinical diagnosis and laboratory diagnosis, was introduced on February 12^16^; Wuhan transportation restrictions were implemented on January 23^15^.

A particularly important challenge is to understand the proportion of transmission that occurs prior to the onset of illness. During the early outbreak, several studies have described the pre-symptomatic transmission of SARS-CoV-2, including a 20-year-old woman from Wuhan believed to have passed on the infection to five of her family members^19^ and a Chinese individual believed to have infected her German business partner^20^, both in the absence of symptoms. The existence of pre-symptomatic transmission indicates that COVID-19 infected individuals can be infectious during the incubation period. However, simple classical susceptible-exposed-infected-recovered (SEIR) models assume weak or no infectiousness during the incubation period^21 14 22^, potentially resulting in an underestimation of overall the infectiousness of COVID-19 cases.

In this study, in order to overcome the difficulties related to describing irregular fluctuations in the transmission dynamics and the limitation of the simple SEIR model, a stochastic susceptible-exposed-infected-quarantined-recovered (SEIQR) model was developed to describe the Wuhan COVID-19 transmission pattern after the initial outbreak stage. This model extends the classic SEIR model by including pre-symptomatic transmission and quarantined status and allows the effects of transportation restrictions and quarantine measures on virus transmission patterns to be estimated while accounting for improvements in the diagnostic capacity over time. After considering varying diagnostic capabilities, we will show that this model can capture the transmission dynamics well and can estimate the reduction in R_e_ precisely.

## Methods

### Data collection

The daily number of new documented COVID-19 cases from January 11 to March 10 in Wuhan, Hubei province, China, were collected from the Wuhan Municipal Health Commission^23^ and the National Health Commission of the People’s Republic of China^24^.

### Description of the SEIQR epidemic model

An SEIQR model was developed to estimate the effect of intervention measures on COVID-19 transmission dynamics in the Wuhan population (Figure 2). In our model, S, E, I, Q and R represent the number of individuals in susceptible, exposed, symptomatically infectious, quarantined, and recovered statuses, with the total population size N = S + E + I + Q + R assumed to be 11 million. Here, we defined susceptible individuals change to exposed individuals after they have had contact with the virus and become infected but not yet symptomatic. Exposed individuals were further divided into two groups: E1, exposed individuals at the latent period who are not able to transmit the disease; E2, exposed individuals not at the latent period who are at a pre-symptomatic stage (referred to pre-symptomatically infectious individuals). The proportions of E1 and E2 out of total exposed individuals were determined using the proportion of the time span of latent period and pre-symptomatic transmission period within the incubation period. In our model, we assumed that all exposed individuals become symptomatic cases after incubation period, and both pre-symptomatically and symptomatically infectious individuals can transmit the disease (Equation 2). For quarantined status, we assumed that only symptomatically infectious individuals can be quarantined. The SEIQR equations were derived as follows:

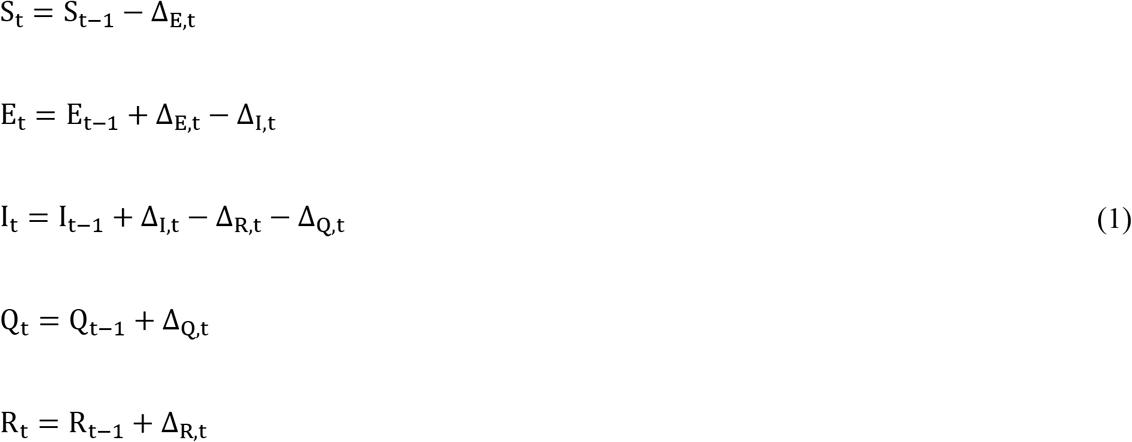

∆_E,t_ is defined as the number of newly exposed individuals before symptom onset, during a time interval from t to t + 1, ∆_I,t_ is the number of newly symptomatically infectious cases (new-onset cases), ∆_Q,t_ is the number of newly quarantined cases, and ∆_R,t_ is the number of newly recovered individuals. We assumed ∆_E,t_, ∆_I,t_, ∆_Q,t_, and ∆_R,t_ follow Poisson distributions:

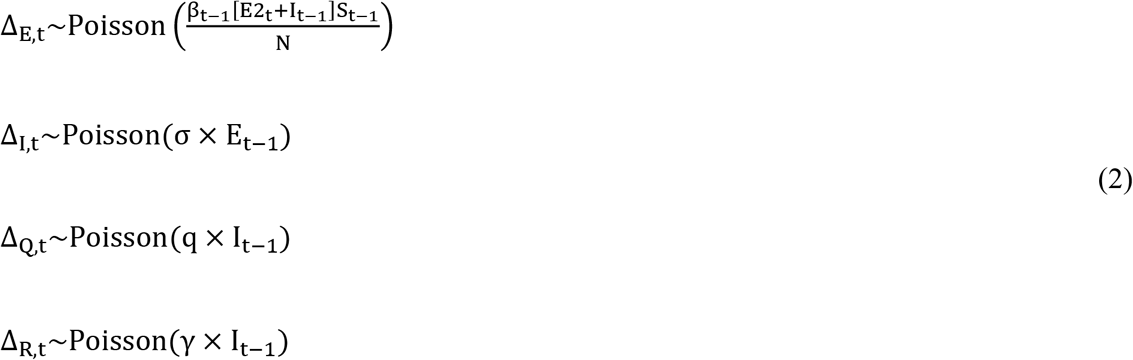

where E2_t_ is the number of pre-symptomatically infectious individuals (E2) at time t, assumed determined as 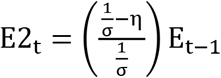, σ is the rate at which exposed individuals become symptomatically infectious cases (1/ σ is the incubation period), η is the latent period, q is the quarantine rate (1/q the time between symptom onset and quarantine start), γ is the recovery rate, expressed by γ = 1/(tau − 1/σ), and tau is the generation time. Here we assumed tau was fixed to be 10 days considering the period from being infected to recovered was generally longer than the observed serial interval (e.g. 7.5 days)^1^ and the infectious period was estimated to be around 10 days by a virology study^25^. Using a constant value of tau can reduce the model uncertainty. β_t_ is the transmission rate on day t. In this model, β_t_ is assumed to be modulated by the Wuhan transportation restriction policy, represented as an exponential relationship with a lag effect:

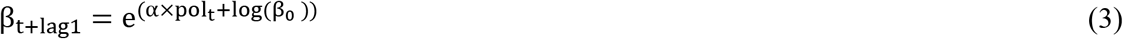

where pol_t_ is an indicator variable for the daily transportation restriction policy, with pol_t_ = 0 if there is no transportation restriction at time t (i.e., before January 23)^15^ and pol_t_ = 1 otherwise. a is the transportation restriction effect coefficient, β_0_ is the basic transmission rate without transportation restrictions, and lag1 indicates the lag time of the transportation restrictions effect on the virus transmission rate assumed to be 6 days^13^. Thus, β _t_ has a constant value throughout the period before the implementation of transportation restriction and change to a different constant value after then.

**Figure 2:**
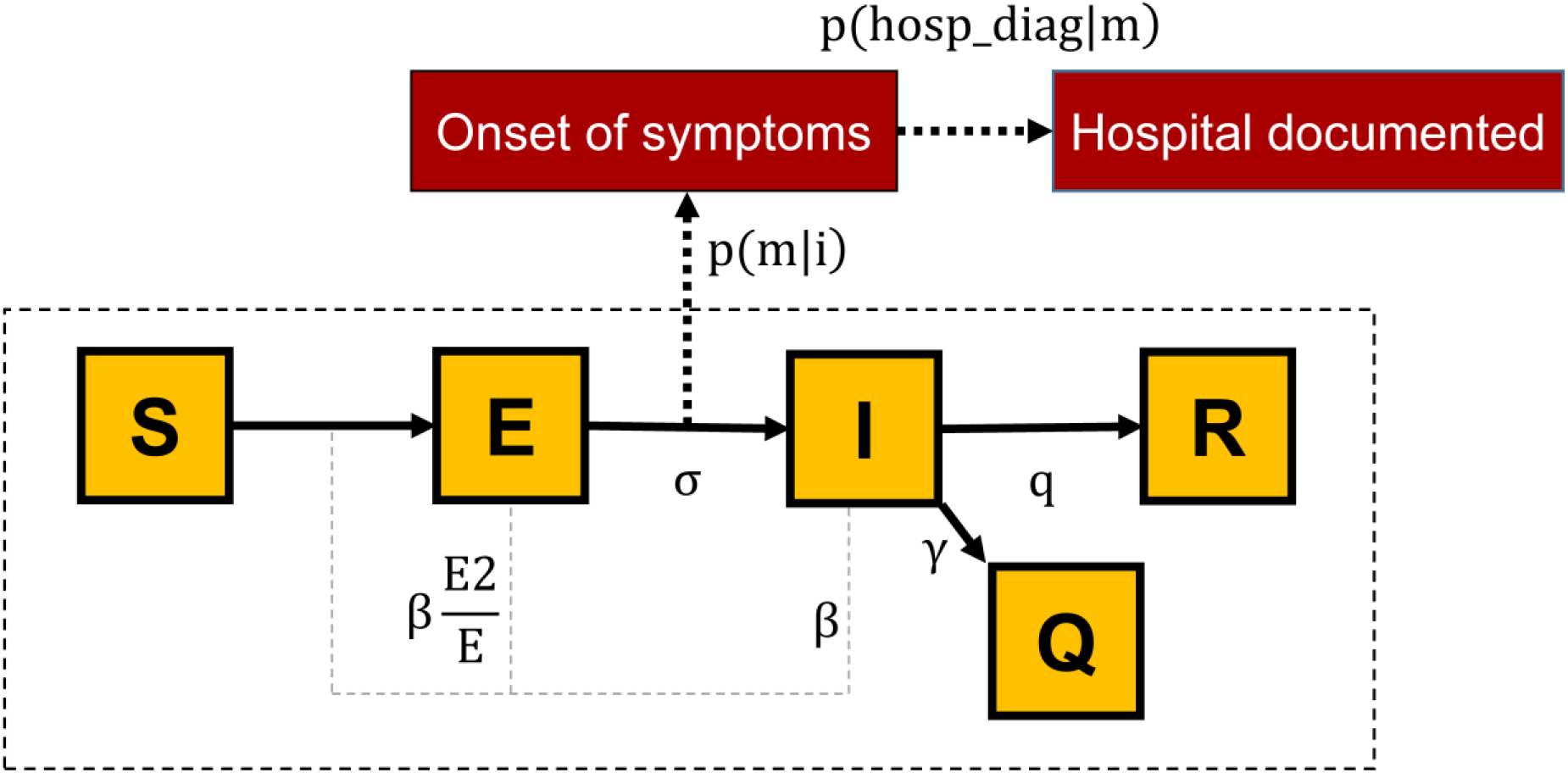
SEIQR model schema. The population is divided into five compartments: S (susceptible), E (exposed), I (symptomatically infectious), Q (quarantined), and R (recovered). E2 is the number of exposed individuals after latent period who are pre-asymptomatically infectious, β is the transmission rate, σ is the incubation rate, q is the quarantine rate, γ is the recovery rate. A fraction of newly symptomatic infections seek for medical care and are eventually documented by hospitals, where p(m|i) is the probability of a symptomatic infectious case seeks medical care, p(hosp_diag|m)_t_ represents the probability that a symptomatic infectious outpatient is diagnosed as COVID-19 case by the hospital.

### Mapping SEIQR model to observed hospital document cases

Model estimates of new-onset cases (∆_I,t_) can not be compared with observed hospital documented cases directly. This is because documented data only captures COVID-19 cases who seek hospital care and are successfully diagnosed, which will only be a proportion of the total number of symptomatically infectious cases in the population estimated in the model. To address this discordance, we introduced an observation model to link the SEIQR model simulated symptomatically infectious cases to the observations. The daily number of hospital documented cases, (hosp_document)_t+lag2_, was assumed followed a normal distribution with the mean defined as the number of newly symptomatically infectious cases ∆_I,t_ that were reported (documented) with the delay lag2 of 6 days^13^:

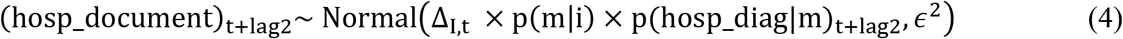

where p(m|i), the probability of a symptomatically infectious case seeks medical care, was assumed to be fixed at 0.8 according to the high motivation of care-seeking behavior in Wuhan^26^. Hospital diagnostic rate, p(hosp_diag|m)_t+lag2_, represents the probability that an infected outpatient is diagnosed as COVID-19 case by the hospital with the delay of lag2. *∊*^2^ is the distribution variance assumed to be 360000. We also defined (prop_doc)_t_, the proportion of documented cases out of total newly symptomatically infectious cases, could be calculated as (prop_doc)_t_ = p(m|i) × p(hosp_diag|m)_t_.

Given that the diagnostic capability progressed over time, hospital diagnostic rate p(hosp_diag|m)_t_ was assumed to have three different values during each of the three periods: p_1_(hosp_diag|m) is the rate for the period prior to January 27 when test kits were limited, p_2_(hosp_diag|m) is the rate for the period between January 27 and Feburary 11 when test kits were sufficient but diagnostic criteria was biased without incorporating clinical diagnosis^7^, and p_3_(hosp_diag|m) is the rate for the period after February 12 when test kits were sufficient and diagnostic criteria became more sensitive based on both clinical diagnosis and laboratory diagnosis^16^. The values of p_1_(hosp_diag|m), p_2_(hosp_diag|m) and p_3_(hosp_diag|m) were estimated after fitting the model to the number of daily hospital documented cases. Hospital documented cases on the specific days of January 27, February 12, and February 13, the dates of change in testing capacity^7 16^(Figure S1), are likely to contain retrospectively documented cases due to the transition to new diagnostic criteria or test kits^27^. Therefore, we removed the original values of these three data and re-filled them by using “na.spline” function in R. That is, the smoothed values of these three dates and the original data of other dates were used during the model fitting process.

### Effective reproductive number R_e_

After obtaining the posterior distributions of model parameters β_t_, σ, q, γ and model status S_t_, the effective reproductive number R_e_(t) before and after the intervention policy was implemented can be calculated using the next-generation matrix approach. Following methods previously described by Diekmann et al.^28^’ the transmission matrices T and Σ can be calculated. Briefly, each element in T represents the average number of newly infected cases in the exposed compartment (E) per unit time due to transmission via a single infected individual in the exposed (E) or infectious group (I), calculated as 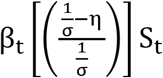 or β_t_S_t_. Σ represents the transitions between model states. R_e_(t) can be calculated as the first eigenvector using the following formula:

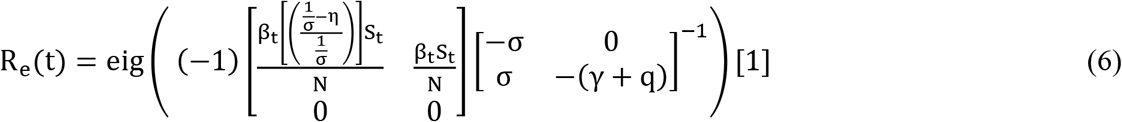

where β_t_, S_t_, σ, q, λ, and N are defined as described above.

### Model-filters and validations

Since the time-varied true number of individuals in S, E, I, Q and R statuses were not directly observable, we used Particle Markov-chain Monte Carlo (PMCMC) method to handle such hidden variables by simultaneously estimating both the parameters and the hidden variables^29^. Our framework of PMCMC contains two parts: the SEIQR transmission model that generates the transmission dynamics and the observation model that maps SEIQR model to observed hospital document cases. All posterior distributions for the epidemiological hidden variables and parameters were obtained using the PMCMC method, implemented in the Nimble R library^30^.

The priors for the parameters were drawn from the following distributions: for the incubation period, 1/σ~U(1,10); for the latent period, η~U(1,7); 1/q~U(1,10), for the time between symptom onset and quarantine start; β_0_~U(0,1) for the basic transmission rate; and α~N(0,1), for transportation control coefficient. In the observation model, the priors for time progressed hospital diagnostic rates were set as uniform distribution: p_1_(hosp_diag|m)/p_2_(hosp_diag|m) ~U(0,1), p_2_(hosp_diag|m)/p_3_(hosp_diag|m) ~U(0,1), p_3_(hosp_diag|m) ~U(0,1).

To assess convergence, three independent chains of the SMC algorithm sets were conducted using 100,000 iterations of 1000 particle samples in each chain. We calculated the effective sample size (ESS) and Gelman-Rubin convergence diagnostic statistics across the three chains.

## Results

### Reconstructing disease dynamics

The daily number of documented COVID-19 cases in Wuhan, increased exponentially up until the first epidemic peak occurring on February 4, and started to fluctuate around the first peak value for about two weeks. Note that the values of the highest peak occurring around the end of the second week in two consecutive days in February were ignored in our study because this peak was primarily caused by the retrospectively documented cases under new diagnostic criteria, whose actual hospitalization date were diversely distributed and can not be traced by our model (Figure S1). The irregular fluctuations can be explained by the effects of interventions and the improved diagnostic capability: the interventions determined the timing of the first peak and may cause a decline pattern afterward; the improved diagnostic capability led to an increase in the number of the documented cases. Together, a high number of cases can be produced for about two weeks. Our stochastic SEIQR model reproduced this irregular pattern by a two-peak dynamic with the first peak occurring on February 4 and the second peak occurring shortly on February 12 (Figure 3). Our estimated times and intensities coincide with the observed epidemic pattern. The estimated incubation period was 5.68 days (95% CI 2.46 - 8.03), and the estimated latent time was 2.82 days (95% CI 1.10 - 5.40) (Table 1).

**Figure 3.**
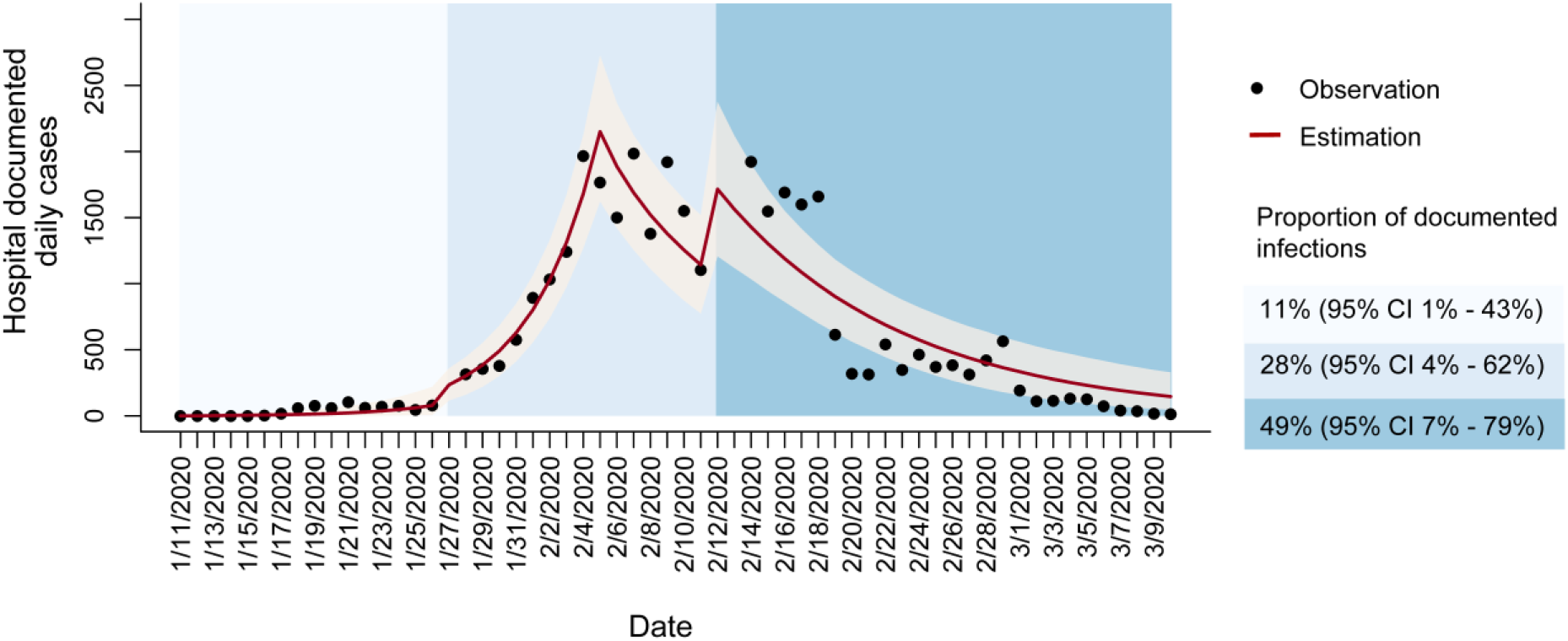
Number of daily hospital documented cases in Wuhan. The red lines represent model-estimated cases, grey shadow represents the 95% prediction interval, black points represent the observed documented cases, blue shaded background denotes incrementally increasing proportions of new documented infections out of total new infections in the corresponding period.

**Table 1.** Parameter estimates of the SEIQR epidemic model. The definitions of the parameters are described. The mean value and 95% credible interval of the posterior distribution of each of the parameters are included. Convergence is diagnosed to have occurred when the value of Gelman-Rubin convergence is close to 1 or the ESS is larger than 200.

| Parameters | Definition | Mean | 95% CI | Gelman-Rubin<br>convergence | ESS |
| --- | --- | --- | --- | --- | --- |
| $1/\sigma$ | Incubation period (days) | 5.68 | (2.46, 8.03) | 1.006 | 261.56 |
| $\eta$ | Latent period (days) | 2.82 | (1.10, 5.40) | 1.005 | 309.46 |
| $1/q$ | Time between symptom<br>onset and quarantine start<br>(days) | 5.44 | (1.99, 9.76) | 1.003 | 477.50 |
| $\alpha$ | Transportation restriction<br>coefficient | -1.96 | (-2.90, -1.21) | 1.003 | 411.77 |
| $\beta_0$ | Basic transmission rate<br>without transportation<br>restrictions | 0.67 | (0.44, 0.97) | 1.001 | 293.01 |
| $p_1(\text{hosp\_diag}\vert \text{m})$ | Hospital diagnostic rate<br>from Jan 11 to Jan 26 | 0.14 | (0.01,0.54) | 1.002 | 396.84 |
| $p_2(\text{hosp\_diag}\vert \text{m})$ | Hospital diagnostic rate<br>from Jan 27 to Feb 11 | 0.35 | (0.05,0.78) | 1.008 | 571.52 |
| $p_3(\text{hosp\_diag}\vert \text{m})$ | Hospital diagnostic rate<br>from Feb 12 to Mar 10 | 0.61 | (0.09,0.98) | 1.004 | 557.22 |

### Effects of intervention measures

Both transportation restrictions and quarantine measures had significant impacts on the effective reproductive number R_e_. The initial value of R_e_ was estimated to be 3.23 (95% CI 2.22 - 4.20) from January 5 to January 28 (Figure 4), but has dropped by 86% to 0.45 (95% CI 0.20 - 0.69) from January 29 to March 4 after the implementation of transportation restrictions, calculated based on the estimated values of transmission rate β_t_ (Figure S2). The estimated time delay to start quarantine after symptom onset was 5.44 days (95% CI 1.99 - 9.76) (Table 1). For limiting the outbreak growth, quarantine measures were important but not essential. Without quarantine measures, the initial value of R_e_ would increase to 4.54 (95% CI 3.65 - 6.79) before the implementation of transportation restrictions, and would become 0.63 (95% CI 0.24 - 1.79) after the implementation of the restrictions (Figure 4). Although R_e_ eventually became less than one, the high initial value of R_e_ would have caused a huge burden of the outbreak. We further tested how the improvements in the diagnostic capacity influenced the estimation of R_e_: about 12%-16% overestimation of R_e_ was found due to a fixed diagnostic capacity (Figure S3); and the model fitting RMSE (root-mean-square error) was increased to be 278.80, comparing to 243.37 from our model, indicating a more accurate prediction was generated from our model taking account of improving diagnostic capabilities.

**Figure 4.**
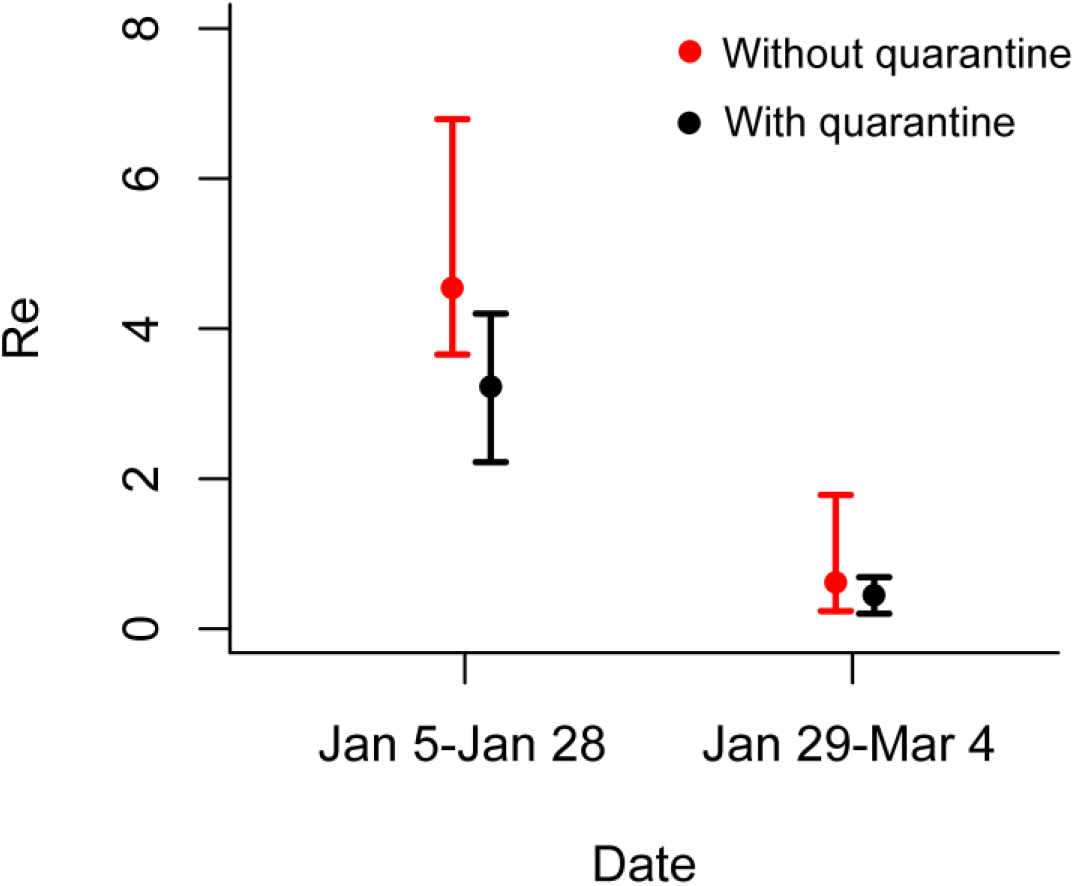
Estimation of the effective reproductive number R_e_ in Wuhan. The red point represents the estimated R_e_ when quarantine measures were not implemented, the black point represents R_e_ when quarantine measures were implemented, and whiskers show the 95% credible intervals.

### Effects of detection capability

During the epidemic, the detection capability of COVID-19 in Wuhan has been improved several times through the increased availability of test kits and the introduction of more sensitive diagnostic criteria (Figure 1). These improvements in the detection capability greatly affected the proportion of documented infections during three periods. From January 11 to January 26, the estimated proportion of documented new infections out of total new infections was 11% (95% CI 1% - 43%), increasing to 28% (95% CI 4% - 62%) following the increase in test kit production on January 26. Then the proportion further raised to 49% (95% CI 7% - 79%) after February 12 when the diagnostic criteria became more sensitive (Figure 5A). The estimated potential cumulative number of infections is correlated with but higher than the observed hospital documented cases in Wuhan, and a sudden surge of hospital documented cases on February 12 can be explained by the introduction of more sensitive diagnostic criteria (Figure 5B).

**Figure 5.**
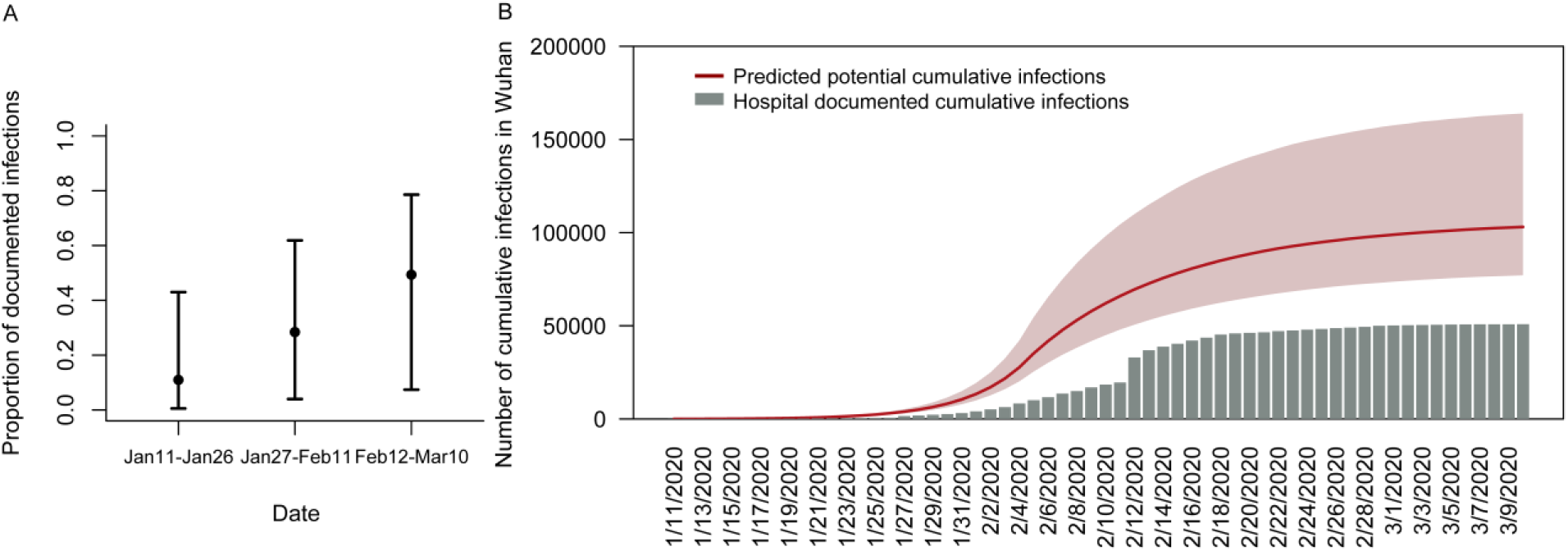
The prediction of temporal diagnostic capability and potential cumulative infections in Wuhan. (A) The estimated proportion of new documented infections out of total new onset infections on different periods with 95% credible intervals. (B) The red line is the predicted potential total cumulative cases, and the red shadow area represents the 95% prediction interval; the grey bar is the hospital documented cumulative cases.

## Discussion

This is the first study to demonstrate the effects of intervention measures on the transmission dynamics in Wuhan while taking account of improvements in diagnostic capacity over time. Our results indicated that the transportation restrictions and quarantine measures together in Wuhan were able to contain local epidemic growth by substantially reducing R_e_ by 86%. This proportion of the reduction in R_e_ was exactly the same as the proportion of the reduction in the average daily number of contacts per person (14.6 vs. 2.0) between a baseline period (before the outbreak) and the outbreak period in a recent study using contact surveys in Wuhan^31^. Since limited studies have estimated the effects of the transportation restrictions in Wuhan, the reduction of contact rate offers valuable information to project the possible effects on the reproduction number. Assuming the transmissibility was proportional to the contact numbers, the reduction ratio of the contact numbers will be proportional to the reduction ratio in R_e_. These results confirm that measuring contact mixing is an accurate way to estimate the impacts of intervention measures. Furthermore, the proportion of undocumented infections was estimated to be reduced during the outbreak, as a consequence of the improvements in diagnostic capability. These findings will help to inform further analysis aimed at developing prevention strategies and evaluating the effects of public health interventions.

While most studies assumed a fixed proportion of documented infections over time, the study presented here estimates an initial proportion of documented infections of 11%, similar to previous predictions of 14% by Ruiyun et al^17^, which progressively increases with the improvement of diagnostic capability. Our results suggest that the increase in the number of cases during the early outbreak needs to be interpreted cautiously, given that the proportion of documented infections is highly dependent on the availability and use of testing kits over time. As detection was enhanced through improved clinical diagnosis^16^, a sharp rise in cumulative cases on February 12 is likely explained by prior onset cases retrospectively documented under new diagnostic criteria. The undocumented infections may be of mild illness or insufficiently serious about seeking treatment^17^. Our results show that the estimated proportion of documented new infections out of total new infections increased to 49% after diagnostic sensitivity was increased.

The estimation of R_e_ in the study from January 5 to January 28 is consistent with other recent studies^32^ (3.11 by Jonathan et al. ^5^, 3.15 by Tian et al. ^21^, 1.4 to 3.9 by Li et al. ^1^). Furthermore, our results demonstrate that both transportation restrictions and quarantine measures were able to reduce COVID-19 transmission. Transportation restrictions, including stopping all forms of public transportation, including trains, and air travel, sharply reduced social contacts thereby reducing virus transmission rates^13 17^. Population behavioral responses (e.g., social distancing, contacts mixing, wearing facemasks, etc.) could change concurrently with the implementation of transportation measures^33 31^. Because a gradual increase in documented hospital cases in February can be partly due to the increased detection capability, the effects of intervention measures (indicated as the reduction in R_e_) was estimated to be larger than previously reported in studies using fixed detection rates over the course of the epidemic. For example, R_e_ was estimated to drop by 55.3% by Kucharski et al^13^. Quarantine of symptomatic infections was also found to be essential in curbing the epidemic. Our model estimated that the time between symptom onset and quarantine start was 5.44 days, similar to the estimates previously reported by Tian et al. (5.19 days)^6^.

The estimated incubation period was 5.68 days which is also consistent with other recent studies^1 21 34 35^. As the estimated latent period is 2.82 days, some transmissions may occur before the symptom onset. Finding ways to reduce possible contact during the pre-symptomatic transmission period may be a critical component in containing the spread of the virus. Given the existence of pre-symptomatic transmission, this study aligns with government recommendations that people who have had close contact with confirmed cases, regardless of whether they show symptoms or not, need to be quarantined for 14 days^36^.

The current study suggests that although intensive transportation restrictions and quarantine measures were critical in containing the COVID-19 outbreak in Wuhan, the improvements in detection capability have to be taken into account in order to evaluate the effectiveness of these intervention measures more accurately. This will allow more meaningful evaluations of public health control effects which will be important for decision in relation to which intervention used in Wuhan should be replicated in other parts of the world in order to effectively control the current pandemic.

## Data Availability

The daily number of new documented COVID-19 cases from January 11 to March 10 in Wuhan, Hubei province, China, were collected from the Wuhan Municipal Health Commission and the National Health Commission of the People's Republic of China.

http://wjw.wuhan.gov.cn/ztzl_28/fk/yqtb/index.shtml

http://www.nhc.gov.cn/xcs/xxgzbd/gzbd_index.shtml

## Acknowledgments

This study was supported by grants from the City University of Hong Kong (#7200573 and #9610416). We thank Dr. Chung Yin (Joey) Leung who has provided invaluable comments. We thank Prof. Mengsu Yang, Prof. Chih-Ching Huang and Prof. Si Zhao Qin at City University of Hong Kong for their suggestions and contributions in the preparation of the manuscript.

## Author contributions

HY and JL designed the research. JL collected the data, carried out the analysis, wrote the first draft. HY, LW and DP critically revised the manuscript, and HY gave final approval for publication.

## Supplementary

**Figure S1.**
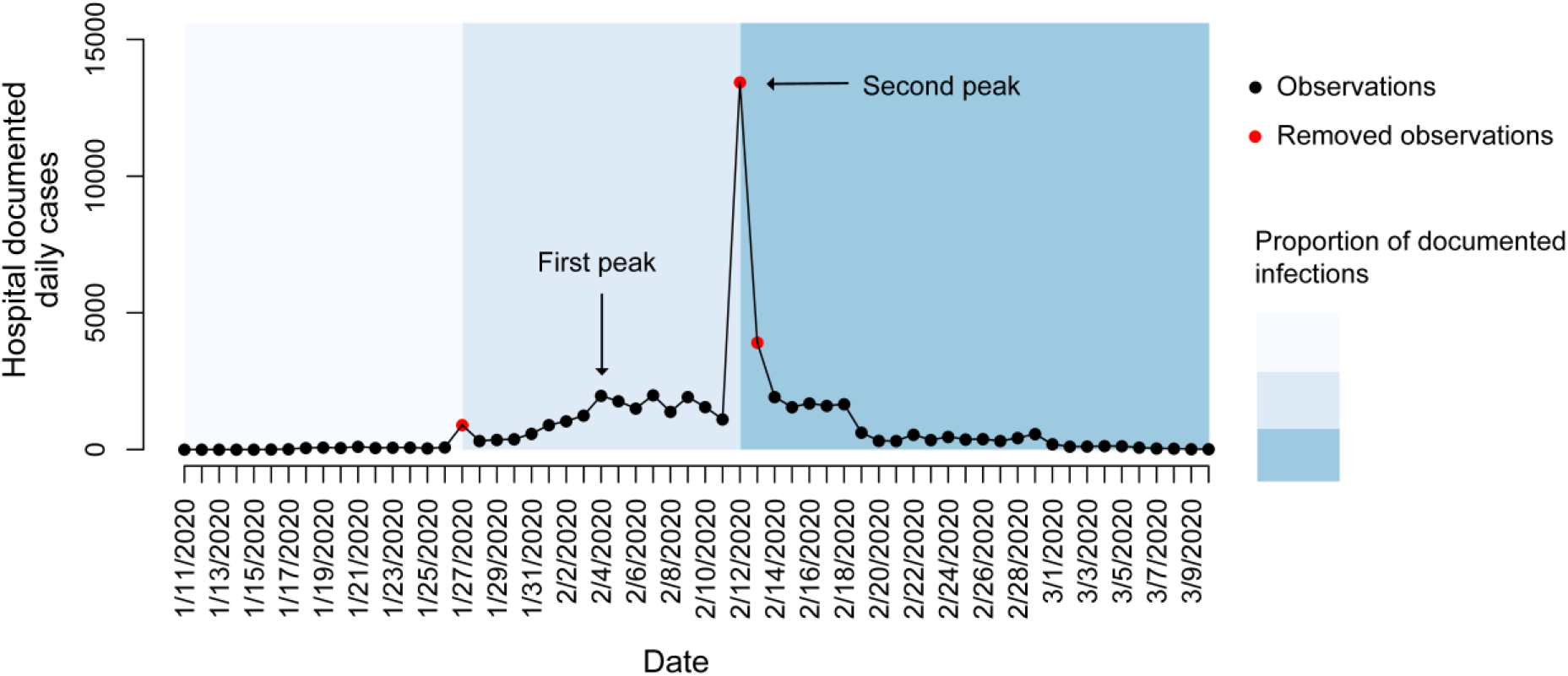
The original observed hospital documented daily cases without removing values. The red points indicate the observed number of cases at the dates when many retrospectively documented cases were counted. Data in these three days were replaced by smoothing values because they contain many retrospectively documented cases. The black points indicate the observed number of cases. Blue shaded background denotes incrementally increasing proportions of new documented infections out of total new infections on the corresponding period caused by improved diagnostic rates.

**Figure S2.**
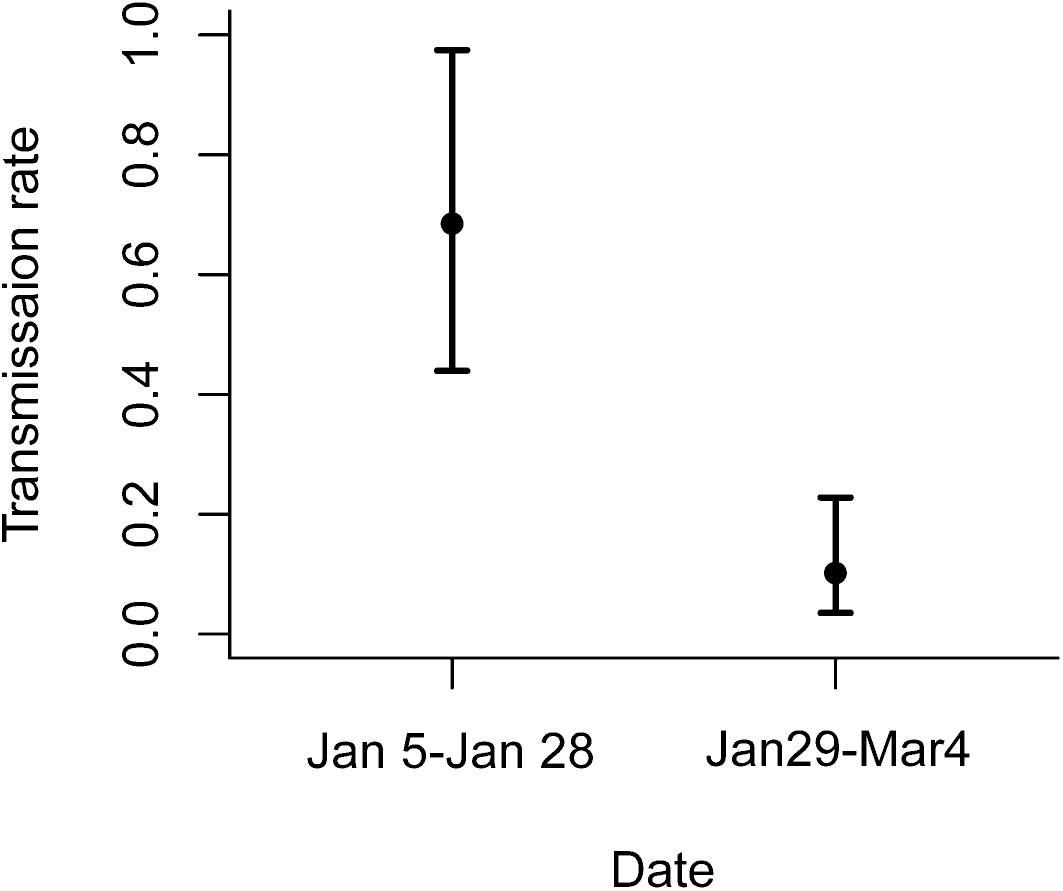
Estimation of the transmission rate β_t_ with 95% credible intervals.

**Figure S3.**
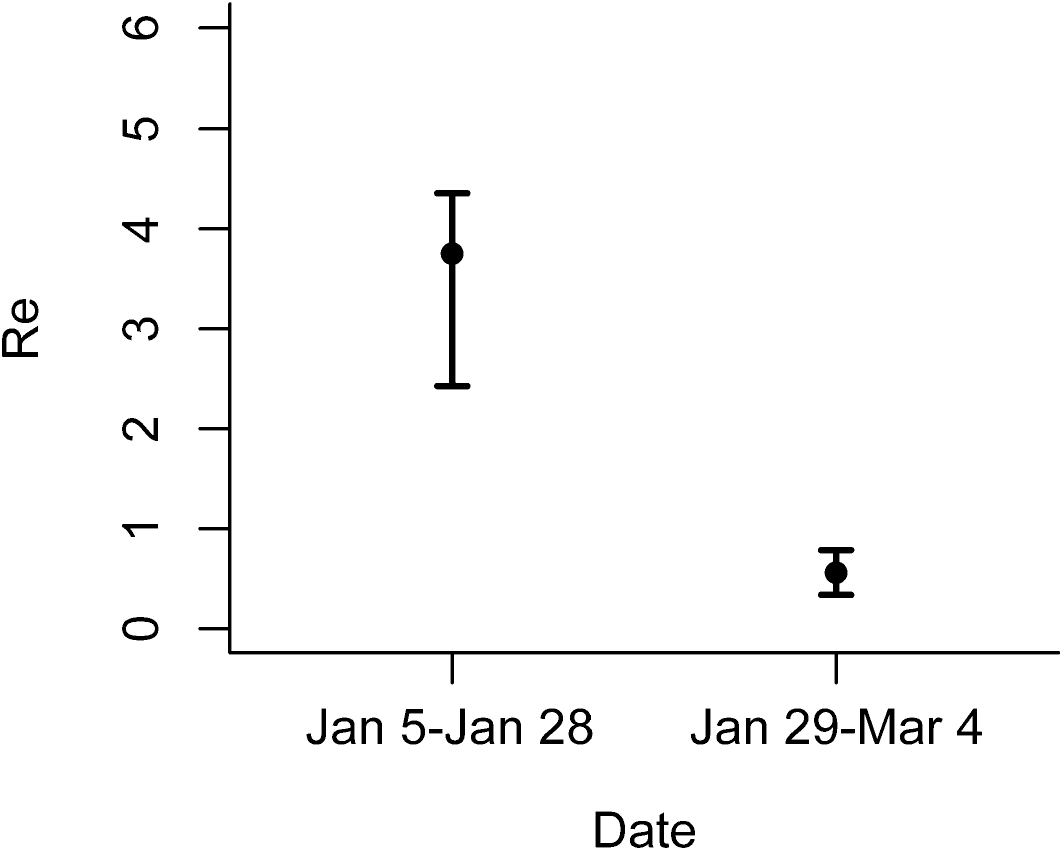
Estimation of the effective reproductive number R_e_ using a fixed hospital diagnostic rate in Wuhan. The fixed hospital diagnostic rate was assumed to be equal to the estimated mean value of the original rate (0.14, see in Table 1) when not considering the improvement of diagnostic capability. R_e_ was estimated to be 3.76 (95% CI 2.43 - 4.36) before the transportation restrictions were implemented and to be 0.56 (95% CI 0.34 - 0.79) after the transportation restrictions were implemented.

**Figure S4.**
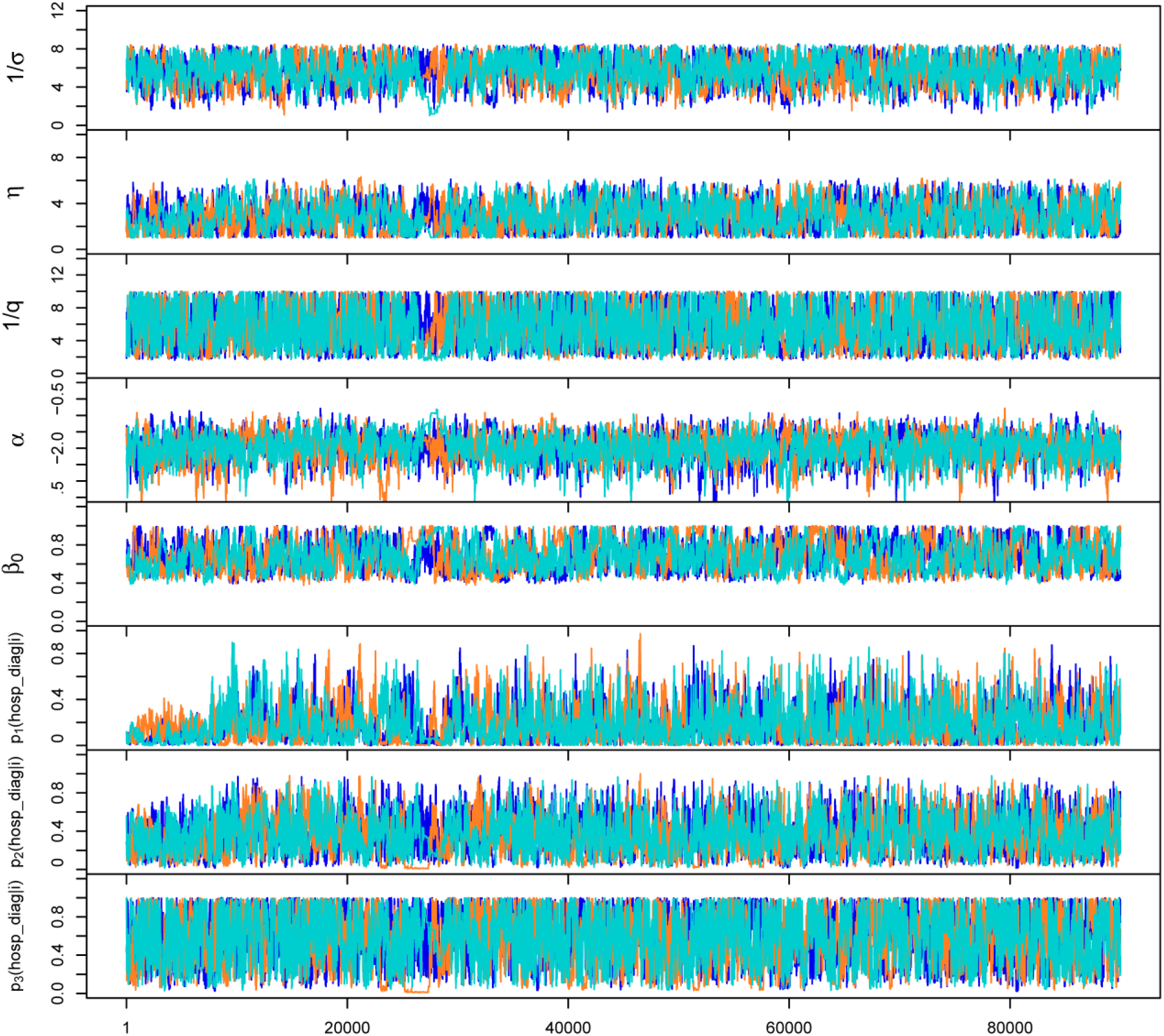
Trace plots of parameter values for the model frame. The three different colours represent three chains.

## Reference

1. Li, Q. et al. Early Transmission Dynamics in Wuhan, China, of Novel Coronavirus–Infected Pneumonia. N. Engl. J. Med. (2020) doi:10.1056/nejmoa2001316.

2. Bedford, J. et al. COVID-19: towards controlling of a pandemic. Lancet 0, (2020).

3. Cobey, S. Modeling infectious disease dynamics. Science (80-.). eabb5659 (2020) doi:10.1126/science.abb5659.

4. Riou, J. & Althaus, C. L. Pattern of early human-to-human transmission of Wuhan 2019 novel coronavirus (2019-nCoV), December 2019 to January 2020. Eurosurveillance 25, 2000058 (20207).

5. Read, J. M., Bridgen, J. R. E., Cummings, D. A. T., Ho, A. & Jewell, C. P. Novel coronavirus 2019-nCoV: early estimation of epidemiological parameters and epidemic predictions. medRxiv (2020) doi:10.1101/2020.01.23.20018549.

6. Tian, H. et al. An investigation of transmission control measures during the first 50 days of the COVID-1777779 epidemic in China. Science (80-.). eabb6105 (2020) doi:10.1126/science.abb6105.

7. Cowling, B. J. et al. Impact assessment of non-pharmaceutical interventions against coronavirus disease 2019 and influenza in Hong Kong: an observational study. Lancet Public Heal. (2020) doi:10.1016/S2468-2667(20)30090-6.

8. Zhao, S. et al. Preliminary estimation of the basic reproduction number of novel coronavirus (2019-nCoV) in China, from 2019 to 2020: A data-driven analysis in the early phase of the outbreak. Int. J. Infect. Dis. 92, 214–217 (2020).

9. Kraemer, M. U. G. et al. The effect of human mobility and control measures on the COVID-19 epidemic in China. Science (80-.). eabb4218 (2020) doi:10.1126/science.abb4218.

10. Chinazzi, M. et al. The effect of travel restrictions on the spread of the 2019 novel coronavirus (COVID-19) outbreak. Science (2020) doi:10.1126/science.aba9757.

11. Du, Z. et al. Risk for Transportation of 2019 Novel Coronavirus Disease from Wuhan to Other Cities in China. Emerg. Infect. Dis. 26, (2020).

12. Zhang, J. et al. Evolving epidemiology and transmission dynamics of coronavirus disease 2019 outside Hubei province, China: a descriptive and modelling study. Lancet Infect. Dis. 3099, 1–10 (2020).

13. Kucharski, A. J. et al. Articles Early dynamics of transmission and control of COVID-19: a mathematical modelling study. Lancet Infect. Dis. (2020) doi:10.1016/S1473-3099(20)30144-4.

14. Lin, Q. et al. A conceptual model for the coronavirus disease 2019 (COVID-19) outbreak in Wuhan, China with individual reaction and governmental action. Int. J. Infect. Dis. 93, 211–216 (2020).

15. The State Council_The People’s Republic of China. http://www.gov.cn/xinwen/2020-01/23/content_5471751.htm.

16. Health Commission of Hubei Province. http://wjw.hubei.gov.cn/bmdt/ztzl/fkxxgzbdgrfyyq/xxfb/202002/t20200213_2025580.shtml.

17. Li, R. et al. Substantial undocumented infection facilitates the rapid dissemination of novel coronavirus (SARS-CoV2). Science (80-.). eabb3221 (2020) doi:10.1126/science.abb3221.

18. SFDA Approves New Coronavirus Nucleic Acid Detection Reagent_Chinese government website. http://www.gov.cn/xinwen/2020-01/27/content_5472368.htm.

19. Bai, Y. et al. Presumed Asymptomatic Carrier Transmission of COVID-19. JAMA (2020) doi:10.1001/jama.2020.2565.

20. Rothe, C. et al. Transmission of 2019-nCoV Infection from an Asymptomatic Contact in Germany. N. Engl. J. Med. (2020) doi:10.1056/nejmc2001468.

21. Tian, H. et al. Early evaluation of the Wuhan City travel restrictions in response to the 2019 novel coronavirus outbreak. medRxiv 2020.01.30.20019844 (2020) doi:10.1101/2020.01.30.20019844.

22. Tang, B. et al. An updated estimation of the risk of transmission of the novel coronavirus (2019-nCov). Infect. Dis. Model. 5, 248–255 (2020).

23. Wuhan Municipal Health Commission. http://wjw.wuhan.gov.cn/ztzl_28/fk/yqtb/index.shtml.

24. National Health Commission of the People’s Republic of China. http://www.nhc.gov.cn/xcs/xxgzbd/gzbd_index.shtml.

25. Zou, L. et al. SARS-CoV-2 viral load in upper respiratory specimens of infected patients. New England Journal of Medicine vol. 382 1177-1179 (2020).

26. The State Council Of The People’s Republic Of China, Policy and regulatory documents; http://www.gov.cn/xinwen/2020-01/24/content_5472017.htm.

27. Tsang, T. K. et al. Effect of changing case definitions for COVID-19 on the epidemic curve and transmission parameters in mainland China: a modelling study. Lancet Public Heal. (2020) doi:10.1016/S2468-2667(20)30089-X.

28. Diekmann, O., Heesterbeek, J. A. P. & Roberts, M. G. The construction of next-generation matrices for compartmental epidemic models. J. R. Soc. Interface 7, 873–885 (2010).

29. Endo, A., van Leeuwen, E. & Baguelin, M. Introduction to particle Markov-chain Monte Carlo for disease dynamics modellers. Epidemics 29, 100363 (2019).

30. NIMBLE – An R package for programming with BUGS models and compiling parts of R. https://r-nimble.org/.

31. Zhang, J. et al. Changes in contact patterns shape the dynamics of the COVID-19 outbreak in China. Science (80-.). eabb8001 (2020) doi:10.1126/science.abb8001.

32. Park, S. W. et al. Reconciling early-outbreak estimates of the basic reproductive number and its uncertainty: framework and applications to the novel coronavirus (SARS-CoV-2) outbreak. doi:10.1101/2020.01.30.20019877.

33. Qian, M. et al. Psychological responses, behavioral changes and public perceptions during the early phase of the COVID-19 outbreak in China: a population based cross-sectional survey. medRxiv 2020.02.18.20024448 (2020) doi:10.1101/2020.02.18.20024448.

34. Backer, J. A., Klinkenberg, D. & Wallinga, J. Incubation period of 2019 novel coronavirus (2019-nCoV) infections among travellers from Wuhan, China, 20–28 January 2020. Eurosurveillance 25, 2000062 (2020).

35. Lauer, S. A. et al. The Incubation Period of Coronavirus Disease 2019 (COVID-19) From Publicly Reported Confirmed Cases: Estimation and Application. Ann. Intern. Med. (2020) doi:10.7326/M20-0504.

36. Jiang, X. et al. Is a 14-day quarantine period optimal for effectively controlling coronavirus disease 2019 (COVID-19)? medRxiv 2020.03.15.20036533 (2020) doi:10.1101/2020.03.15.20036533.

